# First Report of Evaluation of Variant rs11190870 nearby LBX1 Gene with Adolescent Idiopathic Scoliosis Susceptibility in a South-Asian Indian Population

**DOI:** 10.1101/2022.06.28.22276987

**Authors:** Hemender Singh, Shipra, Manish Gupta, Poorvi Bhau, Tania Chalotra, Ruchi Manotra, Nital Gupta, Geetanjali Gupta, Ajay K. Pandita, Mohammad Farooq Butt, Rajesh Sharma, Sarla Pandita, Vinod Singh, Bhavuk Garg, Ekta Rai, Swarkar Sharma

**Affiliations:** Human Genetics Research Group, School of Biotechnology, Shri Mata Vaishno Devi University, Katra, Jammu and Kashmir; Research Associate, Department of Orthopaedics, All India Institute of Medical Sciences, New Delhi; Consultant Orthopedic Surgeon, District Hospital Poonch, Jammu and Kashmir; Radiologist, Department of Radiology, Shri Mata Vaishno Devi Narayana Superspeciality Hospital, Katra, Jammu and Kashmir; Orthopedic Surgeon, Accidental Hospital, Chowki Choura, Jammu, Jammu and Kashmir; Orthopedic Surgeon, Government Medical College, Jammu, Jammu and Kashmir; Radiologist, Professor, Government Medical College, Jammu, Jammu and Kashmir; Radiologist, Chest Disease Hospital, Bakshi Nagar, Jammu, Jammu and Kashmir; Additional Professor, Department of Orthopaedics, All India Institute of Medical Sciences, New Delhi

## Abstract

*LBX1* is a developmental gene involved in skeletal muscle development and somatosensory functioning and proven to be an important gene involved in Adolescent Idiopathic Scoliosis (AIS) etiology. Variant rs11190870 is located 7.5 kb downstream of LBX1 gene and is part of haplotype that is reported to provide risk for AIS. Several studies, including various Genome Wide Association, replication and meta-analyses studies have implicated its association with AIS in different populations. However, any such study is altogether lacking in South-Asian Indian populations. In this first genetic association study for AIS from the region, we tried to replicate association of variant rs11190870 in 95 AIS cases and 282 healthy non-AIS controls from Northwest India. The genotyping was carried out on a Realtime PCR using TaqMan allele discrimination assay and the variant was found to be following Hardy Weinberg equilibrium. The statistical analyses of the genotyping data did not show significant association (p=0.66) of variant rs11190870 with AIS in the population of Northwest India. The results are interesting findings in a population that has never been studied before for AIS susceptibility. However, the findings can be attributed to under power study thus, need evaluation in a large sample set from the population. Interestingly, frequency distribution of the variant in Indian control population datasets was found to be different than other global populations. Linkage Disequilibrium (LD) differences in the genomic region were also observed in these populations while analysing 1000Genomes phase 3 data. It hints at existence of either haplotypic differences in LBX1 locus in South-Asian Indian populations with respect to other populations or genetic heterogeneity in AIS susceptibility. This lays a foundation for genome wide association study (GWAS) in Indian populations cohort, for better understanding of AIS, a task we are pursuing.

## Introduction

Ladybird homeobox 1 (*LBX1)* is one of the important gene that is extensively expressed in skeletal muscles as well as the central nervous system [1]. It plays an important role in the developmental process. It has been reported to be involved in muscle precursor cells migration and development of neural tube [2, 3]. Motor and sensory impairment has been linked to the scoliosis, thus *LBX1* is a great potential candidate for Adolescent Idiopathic Scoliosis (AIS) susceptibility. Various genome-wide association studies [1, 4], replication studies [5-7] as well as meta analyses studies [8-10] have identified *LBX1* as a strong susceptibility locus for AIS in global populations across ethnicities of Non-Hispanic European and East-Asian origins. The variant, rs11190870 is present at 3’ end, 7.5 kb downstream flanking region of *LBX1* gene and reported as variant associated with AIS or part of haplotypes with differential effects in AIS [4]. As rs11190870 lies in close vicinity of *LBX1*, it was proposed to have a regulatory effect on the expression of LBX1 gene, either directly or through other variants in strong LD of it disrupting transcription factor binding sites [11]. This in turn potentially influence downstream molecular mechanisms that contribute to aberrant activity resulting in AIS [12]. All these reasons indicate rs11190870 as a prominent candidate of AIS susceptibility [13] across populations.

In India, the paucity of epidemiological evidences on scoliosis explains the disease’s obscurity and misconceptions. Occurrence and prevalence of AIS in Indian populations has only been documented in Patiala, Punjab (prevalence 0.13%) [14] and Assam (prevalence 0.2%) [15] in literature. Recently, we carried out a study in the Jammu and Kashmir region and observed an overall prevalence of 0.61% with lower female predominance [16]. Further, different association studies and other evidences from various populations worldwide suggest that genetics plays an important role in AIS prevalence. However, any such study is altogether absent in any of the South-Asian Indian populations. With strong evidences that *LBX1* is the prominent candidate gene in AIS susceptibility and variant rs11190870 has been replicated across populations, makes it pertinent to screen it in an Indian population.

Therefore, the overall objective of our present study was to conduct a case-control based association study and screen variant rs11190870, to ascertain its association with AIS, in the Northwest Indian population cohort.

## Methodology

### Sample collection

The study was approved by Shri Mata Vaishno Devi University, Institutional Ethical Review Board (IERB) (IERB Serial No: SMVDU/IERB/18/69 and SMVDU/IERB/18/68). Cases were also collected from collaborators at All India Institute of Medical Sciences, New Delhi, through duly approved study by their ethical committee (Ref No.: IEC-20/08.01.2021, RP-25/2021). 95 AIS cases and 282 healthy controls were included in this study. As only 95 cases could be collected in present study primarily due to low incidence [14-16], 280 controls were estimated (case to control ratio of 1:3) for a power of the study at 80% [based on literature, odds ratio of 1.7 and 55% allelic presence in control set was taken] by Kelsey’s method [17]. 2 ml of venous blood was collected in EDTA vials and kept at -20 degrees till further processing.

### Inclusion of the Cases samples

Majority of the samples were included through school screening programme (details in [16]). These were initially shortlisted using the Adam’s forward bend test and measurement of angle of trunk rotation (ATR). ATR of 7° was considered as the cut off value. It was followed by referral of the individual to the hospitals for clinical examination, radiological examination, exclusion of any other coexisting disorders and confirmation of the condition as AIS. Cobb angle of 10° or more was considered for inclusion of person as a case in the study. Control samples were healthy individuals collected randomly, with age greater than 21 years, from the population of the region.

### Sample processing and DNA isolation

The blood samples were processed with XpressDNA Blood Kit (MagGenome®) for the isolation of genomic DNA. Agarose gel electrophoresis was done to determine the quality of isolated genomic DNA. Quantification of the DNA samples was performed using Nanodrop 2000/2000c (ThermoFisher, USA), followed by the preparation of working dilutions (10 ng/μl).

### Genotyping of variant rs11190870

The Single Nucleotide Polymorphism (SNP) rs11190870 was screened to evaluate its association in studied population group using TaqMan allele discrimination assay conducted on real-time (RT) PCR system (Mx3005P Agilent, USA). TaqMan UNG master mix II (Applied Biosystem, USA) and TaqMan assay (consisted of probe that are VIC and FAM labelled and primers, Applied Biosystem) were used for conducting the genotyping of the variant. According to the manufacturer’s protocol, from 40X stock concentration, 20X TaqMan assay dilution was prepared using TE (Tris-EDTA) buffer. The genotyping of all the samples were performed in 96-well plate which comprised of 93 positive samples and three negative controls (NTC). The total reaction volume was 10 μl each, consisting of TaqMan UNG master mix II (2.5 μl), 20X TaqMan assay (0.25 μl), DNA; 3 μl (10 ng/μl) and water of volume 4.25 μl was added to raise the final reaction to 10μl. The RTPCR conditions comprised of: 95 °C - 10 minutes; 95 °C - 15 seconds and 60 °C - 1 minute for 40 cycles. The post PCR detection system measured the allele-specific fluorescence, and automatically the calling of alleles was done. Samples with failure calls were repeated for genotyping. For QC, 10% of the genotyped samples were selected randomly for re-genotyping and calls were observed with 100% concordance.

### Data analyses

Statistical analyses of the genotyping data was performed after the completion of experimental work using Plink v1.09. Genotypic and allele frequencies were estimated and also evaluated for Hardy Weinberg equilibrium (HWE). Chi Square test was employed to 2×2 and 2×3 tables to assess significance of differences in allelic and genotypic distributions, respectively. The association of SNP with AIS was evaluated by calculating odds ratio (OR), 95% confidence interval (CI) along with significance level (p value). Frequency and Genotype data for the variant rs11190870 in various population groups was retrieved from 1000GENOMES:phase 3 through Ensembl (www.ensembl.org) and IndiGenomes (https://clingen.igib.res.in/indigen/) for comparison (details in Table 1). Linkage Disequilibrium (LD) measures and plots w.r.t variant were also collected for comparison from Ensembl based on 1000GENOMES:phase 3 data for global populations: CEU (Utah Residents with Northern and Western European ancestry), PEL (Peruvian in Lima, Peru), YRI (Yoruba in Ibadan, Nigeria), CHB (Han Chinese in Bejing, China), JPT (Japanese in Tokyo, Japan), and populations of South-Asian Indian origin: GIH (Gujarati Indian in Houston, TX), BEB (Bengali in Bangladesh), ITU (Indian Telugu in the UK), PJL (Punjabi in Lahore, Pakistan).

**Table 1:**
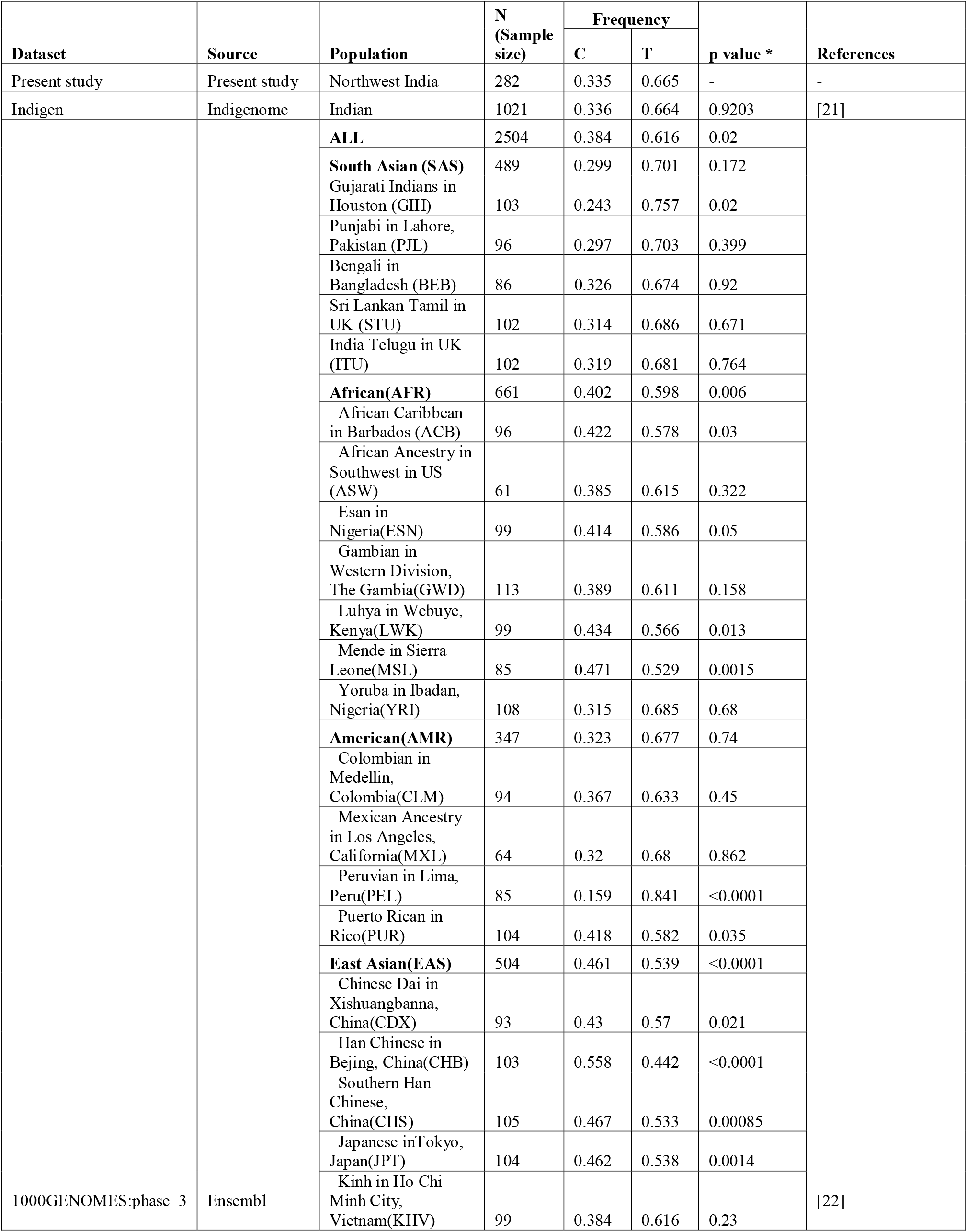
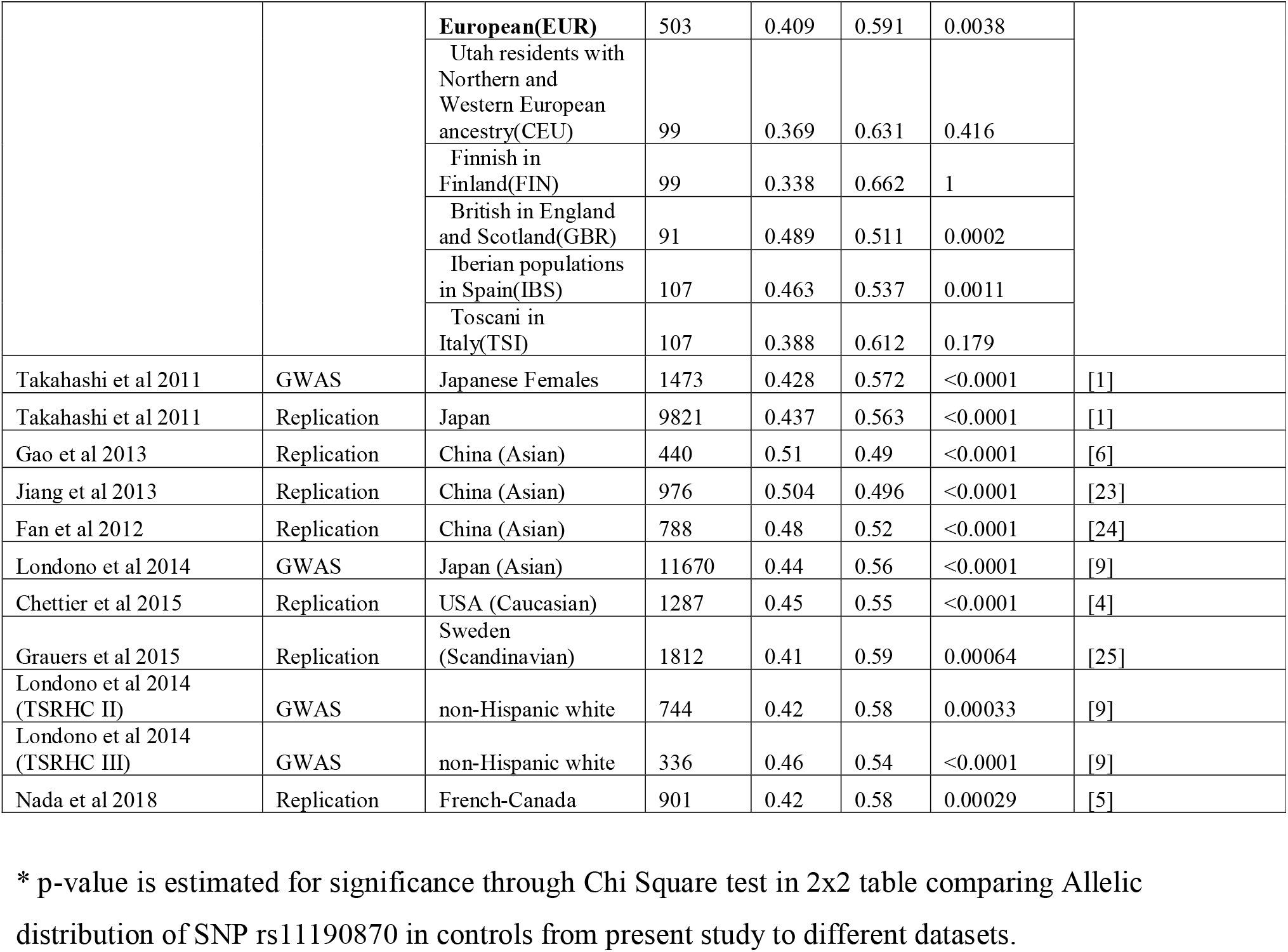
Allele frequency distribution of SNP rs11190870 in healthy South-Asian Indian datasets and other global ethnic population groups.

## Results and Discussion

AIS being one of the frequent complications of the musculoskeletal system has been explored for its genetic susceptibility across populations using various study designs [18]. Several genes and their variants were identified for their association with AIS. One such variant, rs11190870, which is located near *LBX1* gene was observed as most widely associated variant with AIS, especially in populations of Non-Hispanic European and East-Asian origins [9, 19, 20].

In this first replication study, we examined the association of variant rs11190870 near *LBX1* with AIS predisposition in Northwest Indian population. The genotypes distributions, in cases (CC = 0.137, CT = 0.358 and TT = 0.505) and controls (CC = 0.135, CT = 0.397, TT = 0.468) separately as well as in total samples, were found to be following HWE. The distribution of allele frequencies also did not show significant difference in cases (C = 0.316, T = 0.684) and controls (C = 0.333, T = 0.667) and variant was not found associated (p = 0.66) with AIS in present population cohort. As a result of near similar distribution of alleles in both cases and controls led to the non-association of the variant in studied population group. Despite the fact that the present work was planned with 80% power of the study, due to observation of increased frequency of risk allele (T = 0.67) in present control population group, this study (95 cases and 282 controls) has been found at power of 49%. To increase the power of the study, we also compared the genotypes distribution of our cases with publicly available control dataset, “IndiGenomes” (n=1021, case to control ratio 1:10.75) of South-Asian Indian Population, [21], The allelic distribution (C = 0.336, T = 0.664) and results remained near same with power of the study at 56%.

Interestingly, it was also observed that “T” allele (ancestral and AIS risk allele of variant rs11190870) was found to be in relatively lower frequencies in controls from populations of Non-Hispanic European and East-Asian origins either in various studies or in 1000GENOMES:phase 3 data (Table 1). Similar observation was made with respect to populations of African origins, other than YRI (Yoruba in Ibadan, Nigeria) population where T allele was observed with higher frequency (Table 1). “T” allele frequency, in controls from present study as well as healthy individuals in other public datasets of populations of South-Asian Indian origin, also showed higher values (Table 1) indicating the derived allele of variant (“C” allele) is less frequent in the populations. Populations of Hispanic and other American origins also showed almost similar allelic distribution of variant rs11190870 as of South-Asian Indian origin populations (Table 1). Thus, it would be interesting to see association status of the variant in Hispanic and other American origins populations for AIS susceptibility.

We also compared LD plots for variations that showed high LD (r^2^ >0.8) w.r.t variant rs11190870 in various reference populations datasets from 1000GENOMES:phase 3 data (Supplementary Figure 1) as well as tabulated these variations through Ensembl (Supplementary File). Differences in LD were observed across different populations as can be visualised and noted in tables (Supplementary Figure 1 and Supplementary File).

Moreover, the ancestral allele (“T” allele) is reported as the risk allele, indicating the SNP rs11190870 is probably the proxy SNP for the risk signal and actual casual variant/s might be different but in strong LD to rs11190870 in populations of Non-Hispanic European and East-Asian origins, as explained in Figure 1. Further, the casual variant/s appears to occur in ancestral state of rs11190870. Thus, in populations where LD is not maintained, or have lower frequencies of casual variant/s (Figure 1), the variant rs11190870 may not show association, despite potential role of gene in etiology of the disease in the respective population.

**Figure 1:**
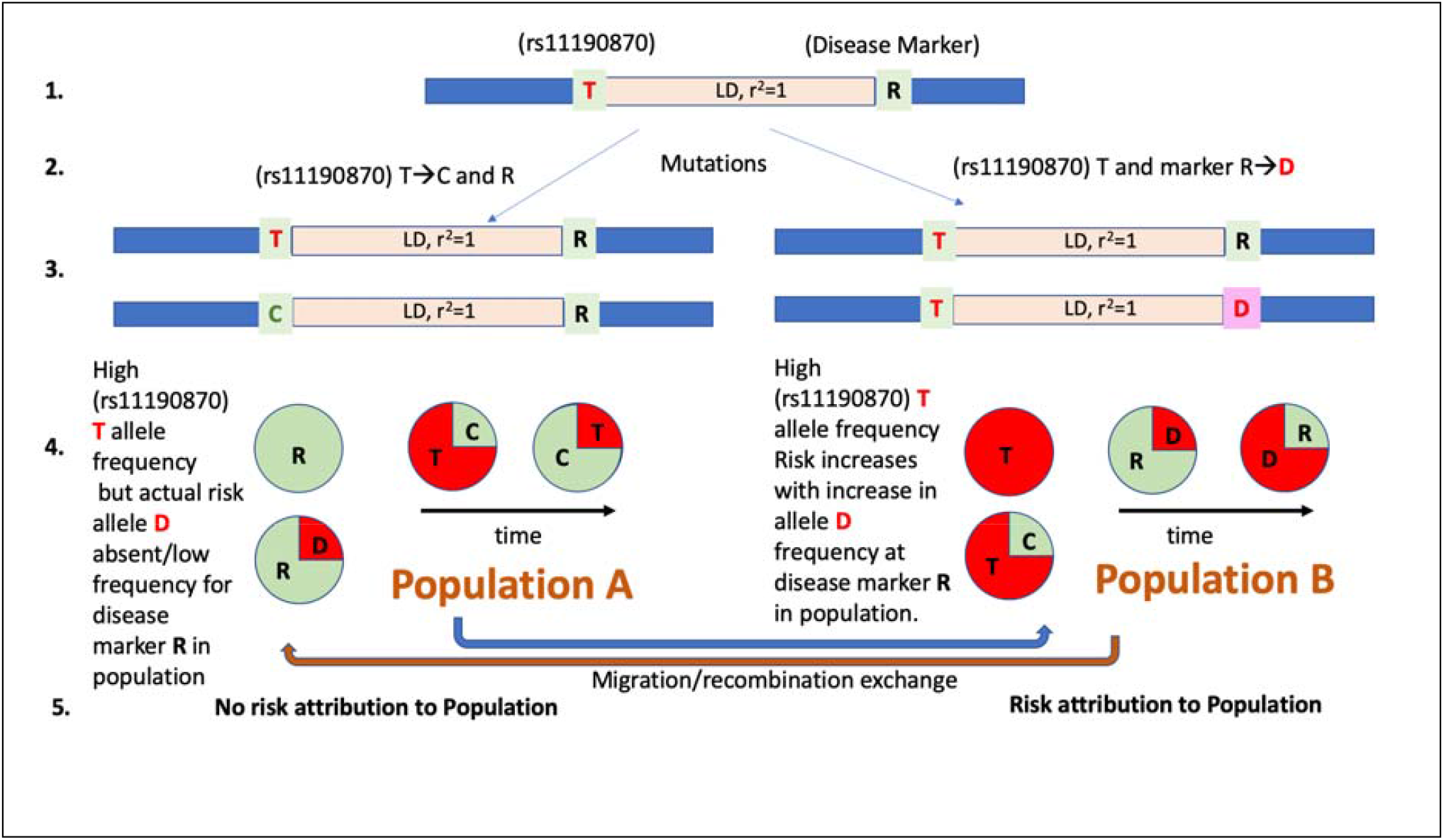
Representation of potential mechanism of association of ancestral allele of the variant rs11190870 with AIS and differential outcomes in different populations. 1. “T” allele is the ancestral state of the variant rs11190870 present in absence of T→C mutation. “R” denotes ancestral state of casual disease marker and in strong LD with rs11190870. 2. Mutations T→C at variant rs11190870 occurs at any point of time in population A whereas ancestral state R remains at disease marker locus. Independently, R→D occurs at any point of time in another population but with ancestral state “T” at variant rs11190870. 3. It results in Haplotype T-R and C-R at rs11190870 - disease marker, in the Population A whereas, Haplotype T-R and T-D at rs11190870 - disease marker, in Population B 4. Population expansion occurs resulting in increase of “C” allele at variant rs11190870 in population A and “R” status at disease locus. However in Population B, risk allele “D” frequency increases with time as population expands. Also, chance of migration and recombination may introduce “C” allele in population B and “D” allele in population A. 5. Thus, despite high allele frequency of “T” allele in population A, it may not indicate disease risk associated with it in population A.

## Conclusion

This is the first genetic study that provides an insight in AIS predisposition in a South Asian Indian population. It reports non-association of rs11190870 in Northwest Indian population cohort, warranted to be replicated with increased sample size. The present study also highlights the need for screening of other SNPs across the LBX1 gene for association. It also highlights the possibility of the potential haplotypic differences or genetic heterogeneity and lays a foundation for carrying out a genome wide association study in South Asian Indian populations for better understanding of AIS.

## Data Availability

All data produced in the present work are contained in the manuscript.

## Acknowledgment

All the authors acknowledge Dr. Varun Sharma, Dr. Indu Sharma, Dr. Ankit Mahajan, Akash Sharma, Sonakshi Modeel, Hetender Singh, NSS SMVDU Volunteers and M.Sc. Dissertation students at SMVDU, International Volunteer Foundation, Jammu and Indian Red Cross Society, District Branch Jammu for their assistance in population screening and sample collection. Authors acknowledge Prof. Manoj Kumar Dhar, School of Biotechnology, University of Jammu, and Prof. Sanjana Kaul, School of Biotechnology, University of Jammu for facilitating in quantification of DNA samples. Authors acknowledge Central Massarray Facility, SoBT SMVDU for facilitation of Genotyping of the samples.

## Author Contributions

SS, ER conceived the study and VS assisted in study design and execution. HS, S, PB, TC and RM carried out population screening, sample collection, DNA isolation and genotyping of the samples. HS, S performed statistical analyses of the data. HS wrote the manuscript. MG, NG, GG, AKP, MFB, RS, SP, BG are clinical collaborators who provided clinical evaluation of the cases and samples. SS, ER, VS critically reviewed the manuscript.

## Competing Interests

The authors declare no competing interests associated with MS.

## Figure Legends

**Supplementary Figure 1:**
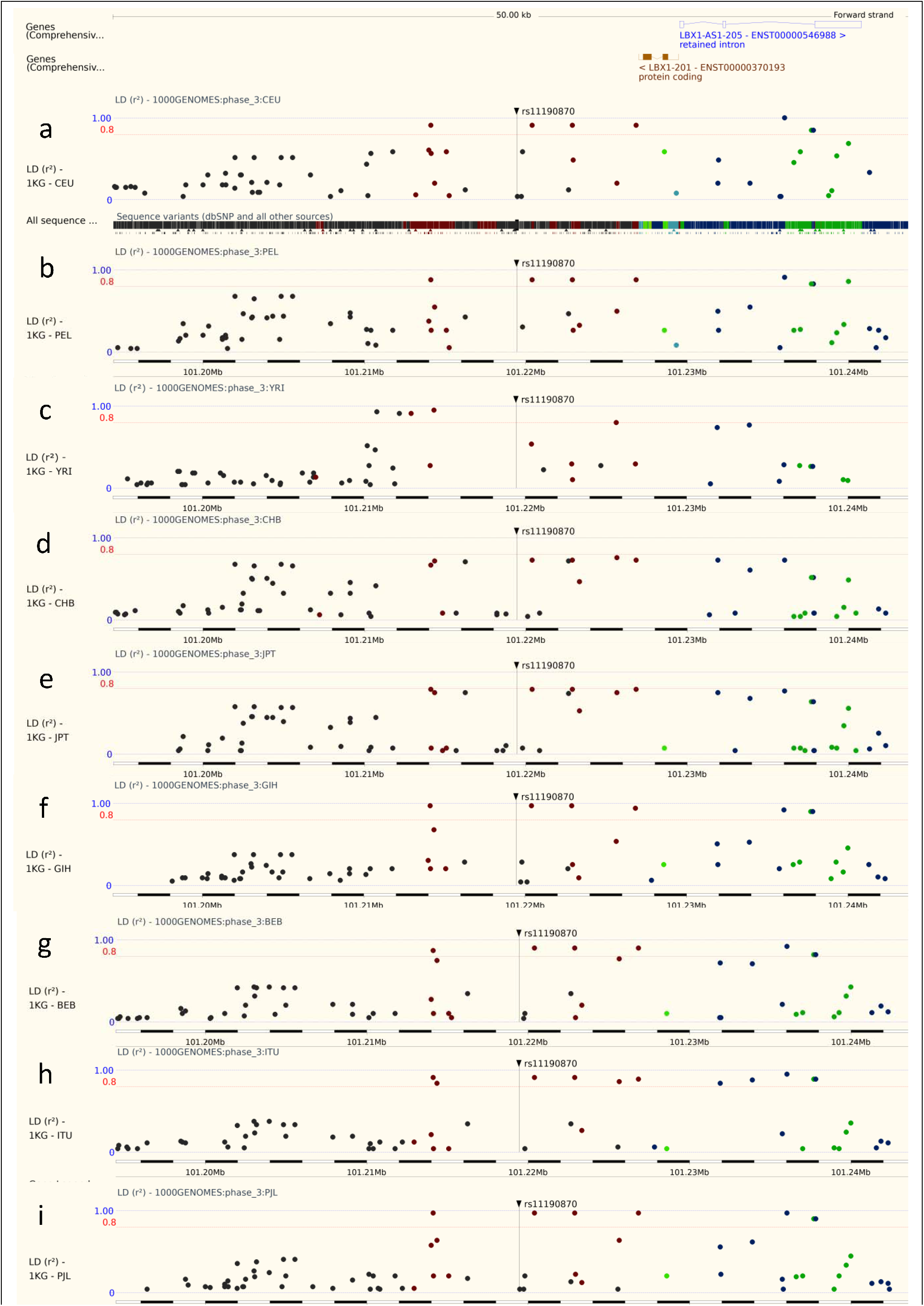
LD Plots of SNPs based on r^2^ value in ∼50kb of genomic region of rs11190870 in different reference populations of the world from 1000Genomes:phase 3 data. a. CEU (Utah Residents with Northern and Western European ancestry), b. PEL (Peruvian in Lima, Peru), c. YRI (Yoruba in Ibadan, Nigeria), d. CHB (Han Chinese in Bejing, China), e. JPT (Japanese in Tokyo, Japan), f. GIH (Gujarati Indian in Houston, TX), g. BEB (Bengali in Bangladesh), h. ITU (Indian Telugu in the UK), i. PJL (Punjabi in Lahore, Pakistan)

**Supplementary File: List of variations linked to rs11190870 with LD (r2 >0.8) in different reference populations sets from 1000GENOMES:phase 3**

## Notes

### Competing Interest Statement

The authors have declared no competing interest.

### Funding Statement

This study did not receive any funding

### Author Declarations

Ethics committee of Shri Mata Vaishno Devi University [Institutional Ethical Review Board (IERB) (IERB Serial No: SMVDU/IERB/18/69 and SMVDU/IERB/18/68)]and Ethics committee of All India Institute of Medical Sciences, New Delhi [Institute ethics committee (IEC), All India Institute of Medical Sciences (Ref No.: IEC-20/08.01.2021, RP-25/2021)] gave approval for this work.

