## Supplementary File for "First Report of Evaluation of Variant rs11190870 nearby LBX1 Gene with Adolescent Idiopathic Scoliosis Susceptibility in a South-Asian Indian Population"

### Supplementary file (source: www.ensembl.org)

### Variations linked to rs11190870 in 1000GENOMES:phase 3 populations with LD (r2 >0.8)

Reference:
Cunningham, F., J. E. Allen, J. Allen, J. Alvarez-Jarreta, M R. Amode, Irina M. Armean, O. Austine-Orimoloye, Andrey G. Azov, I. Barnes, R. Bennett, A. Berry, J. Bhai, A. Bignell, K. Billis, S. Boddu, L. Brooks, M. Charkhchi, C. Cummins, L. Da Rin Fioretto, C. Davidson, K. Dodiya, S. Donaldson, B. El Houdaigui, T. El Naboulsi, R. Fatima, C. G. Giron, T. Genez, Jose G. Martinez, C. Guijarro-Clarke, A. Gymer, M. Hardy, Z. Hollis, T. Hourlier, T. Hunt, T. Juettemann, V. Kaikala, M. Kay, I. Lavidas, T. Le, D. Lemos, J. C. Marugán, S. Mohanan, A. Mushtaq, M. Naven, Denye N. Ogeh, A. Parker, A. Parton, M. Perry, I. Piližota, I. Prosovetskaia, Manoj P. Sakthivel, Ahamed Imran A. Salam, Bianca M. Schmitt, H. Schuilenburg, D. Sheppard, José G. Pérez-Silva, W. Stark, E. Steed, K. Sutinen, R. Sukumaran, D. Sumathipala, M.-M. Suner, M. Szpak, A. Thormann, F. F. Tricomi, D. Urbina-Gómez, A. Veidenberg, Thomas A. Walsh, B. Walts, N. Willhoft, A. Winterbottom, E. Wass, M. Chakiachvili, B. Flint, A. Frankish, S. Giorgetti, L. Haggerty, Sarah E. Hunt, Garth R. IIsley, Jane E. Loveland, Fergal J. Martin, B. Moore, Jonathan M. Mudge, M. Muffato, E. Perry, M. Ruffier, J. Tate, D. Thybert, Stephen J. Trevanion, S. Dyer, Peter W. Harrison, Kevin L. Howe, Andrew D. Yates, Daniel R. Zerbino and P. Flicek (2021). "Ensembl 2022." Nucleic Acids Research 50(D1): D988-D995.

#### 1000GENOMES:phase 3 through Ensembl (www.ensembl.org)

##### rs11190870 SNP

| | |
|---|---|
| **Most severe consequence** | **intergenic variant** |
| **Alleles** | **T/A/C** \| Ancestral: **T** \| MAF: **0.38** (C) \| Highest population MAF: **0.49** |
| **Change tolerance** | CADD: A:22.4, C:22.5 \| GERP: 2.59 |

#### Linkage disequilibrium

##### Pairwise linkage disequilibrium data by population

##### Links to linkage disequilibrium data by population

Show All entries — Show/hide columns — Filter

| Population | Description | LD Manhattan plot | Variants in high LD | LD plot | |
|---|---|---|---|---|---|
| **African** | | | | | |
| 1000GENOMES:phase_3:**ACB** | African Caribbean in Barbados | View plot | Show | View plot | View table |
| 1000GENOMES:phase_3:**ASW** | African Ancestry in Southwest US | View plot | Hide | View plot | View table |
| 1000GENOMES:phase_3:**ESN** | Esan in Nigeria | View plot | Show | View plot | View table |
| 1000GENOMES:phase_3:**GWD** | Gambian in Western Division, The... (more) | View plot | Show | View plot | View table |
| 1000GENOMES:phase_3:**LWK** | Luhya in Webuye, Kenya | View plot | Hide | View plot | View table |
| 1000GENOMES:phase_3:**MSL** | Mende in Sierra Leone | View plot | Show | View plot | View table |
| 1000GENOMES:phase_3:**YRI** | Yoruba in Ibadan, Nigeria | View plot | Hide | View plot | View table |
| **American** | | | | | |
| 1000GENOMES:phase_3:**CLM** | Colombian in Medellin, Colombia | View plot | Show | View plot | View table |
| 1000GENOMES:phase_3:**MXL** | Mexican Ancestry in Los Angeles... (more) | View plot | Show | View plot | View table |
| 1000GENOMES:phase_3:**PEL** | Peruvian in Lima, Peru | View plot | Show | View plot | View table |
| 1000GENOMES:phase_3:**PUR** | Puerto Rican in Puerto Rico | View plot | Show | View plot | View table |
| **East Asian** | | | | | |
| 1000GENOMES:phase_3:**CDX** | Chinese Dai in Xishuangbanna, China | View plot | Hide | View plot | View table |
| 1000GENOMES:phase_3:**CHB** | Han Chinese in Bejing, China | View plot | Hide | View plot | View table |
| 1000GENOMES:phase_3:**CHS** | Southern Han Chinese, China | View plot | Hide | View plot | View table |
| 1000GENOMES:phase_3:**JPT** | Japanese in Tokyo, Japan | View plot | Hide | View plot | View table |
| 1000GENOMES:phase_3:**KHV** | Kinh in Ho Chi Minh City, Vietnam | View plot | Hide | View plot | View table |
| **European** | | | | | |
| 1000GENOMES:phase_3:**CEU** | Utah residents with Northern and... (more) | View plot | Hide | View plot | View table |
| 1000GENOMES:phase_3:**FIN** | Finnish in Finland | View plot | Show | View plot | View table |
| 1000GENOMES:phase_3:**GBR** | British in England and Scotland | View plot | Show | View plot | View table |
| 1000GENOMES:phase_3:**IBS** | Iberian populations in Spain | View plot | Show | View plot | View table |
| 1000GENOMES:phase_3:**TSI** | Toscani in Italy | View plot | Show | View plot | View table |
| **South Asian** | | | | | |
| 1000GENOMES:phase_3:**BEB** | Bengali in Bangladesh | View plot | Show | View plot | View table |
| 1000GENOMES:phase_3:**GIH** | Gujarati Indian in Houston, TX | View plot | Show | View plot | View table |
| 1000GENOMES:phase_3:**ITU** | Indian Telugu in the UK | View plot | Hide | View plot | View table |
| 1000GENOMES:phase_3:**PJL** | Punjabi in Lahore, Pakistan | View plot | Hide | View plot | View table |
| 1000GENOMES:phase_3:**STU** | Sri Lankan Tamil in the UK | View plot | Show | View plot | View table |
| **GGVP:ALL** | **All populations from the Gambian...** (more) | | | | |
| GGVP:**GWD** | Population from the Gambian Genome... (more) | View plot | Show | View plot | View table |
| GGVP:**GWF** | Population from the Gambian Genome... (more) | View plot | Show | View plot | View table |
| GGVP:**GWJ** | Population from the Gambian Genome... (more) | View plot | Show | View plot | View table |
| GGVP:**GWW** | Population from the Gambian Genome... (more) | View plot | Show | View plot | View table |

| Variant | Location | Distance (bp) | $r^2$ | D' | Associated phenotype(s) | Consequence Type | Located in gene(s) | Gene phenotype(s) |
|---|---|---|---|---|---|---|---|---|
| rs11598177 | 10:101220399 | 949 | 0.975 | 1.000 | - | regulatory region variant | - | - |
| rs1322332 | 10:101222891 | 3441 | 0.975 | 1.000 | - | regulatory region variant | - | - |
| rs1535462 | 10:101214115 | 5335 | 0.975 | 1.000 | - | TF binding site | - | - |
| rs1322331 | 10:101226832 | 7382 | 0.975 | 1.000 | - | regulatory region variant | - | - |
| rs594791 | 10:101236039 | 16589 | 0.975 | 1.000 | - | intron variant | LBX1-AS1 | - |
| rs679206 | 10:101237693 | 18243 | 0.901 | 0.974 | - | non coding transcript exon variant | LBX1-AS1 | - |
| rs678741 | 10:101237824 | 18374 | 0.901 | 0.974 | - | intron variant | LBX1-AS1 | - |

##### Variants linked to rs11190870 in 1000GENOMES:phase_3:ITU

[back to top]

Show All entries Show/hide columns Filter

| Variant | Location | Distance (bp) | $r^2$ | D' | Associated phenotype(s) | Consequence Type | Located in gene(s) | Gene phenotype(s) |
|---|---|---|---|---|---|---|---|---|
| rs594791 | 10:101236039 | 16589 | 0.956 | 1.000 | - | intron variant | LBX1-AS1 | - |
| rs11598177 | 10:101220399 | 949 | 0.915 | 1.000 | - | regulatory region variant | - | - |
| rs1322332 | 10:101222891 | 3441 | 0.915 | 1.000 | - | regulatory region variant | - | - |
| rs1535462 | 10:101214115 | 5335 | 0.912 | 0.977 | - | TF binding site | - | - |
| rs1322331 | 10:101226832 | 7382 | 0.895 | 1.000 | - | regulatory region variant | - | - |
| rs679206 | 10:101237693 | 18243 | 0.895 | 1.000 | - | non coding transcript exon variant | LBX1-AS1 | - |
| rs678741 | 10:101237824 | 18374 | 0.895 | 1.000 | - | intron variant | LBX1-AS1 | - |
| rs625039 | 10:101233892 | 14442 | 0.889 | 0.976 | - | intron variant | LBX1-AS1 | - |
| rs1407409 | 10:101225650 | 6200 | 0.868 | 0.975 | - | regulatory region variant | - | - |
| rs3950032 | 10:101214352 | 5098 | 0.847 | 0.975 | - | TF binding site | - | - |
| rs1322330 | 10:101231902 | 12452 | 0.845 | 0.930 | - | intron variant | LBX1-AS1 | - |

##### Variants linked to rs11190870 in 1000GENOMES:phase_3:CEU

[back to top]

Show/hide columns Filter

| Variant | Location | Distance (bp) | $r^2$ | D' | Associated phenotype(s) | Consequence Type | Located in gene(s) | Gene phenotype(s) |
|---|---|---|---|---|---|---|---|---|
| rs594791 | 10:101236039 | 16589 | 1.000 | 1.000 | - | intron variant | LBX1-AS1 | - |
| rs11598177 | 10:101220399 | 949 | 0.918 | 1.000 | - | regulatory region variant | - | - |
| rs1322332 | 10:101222891 | 3441 | 0.918 | 1.000 | - | regulatory region variant | - | - |
| rs1535462 | 10:101214115 | 5335 | 0.918 | 1.000 | - | TF binding site | - | - |
| rs1322331 | 10:101226832 | 7382 | 0.918 | 1.000 | - | regulatory region variant | - | - |
| rs679206 | 10:101237693 | 18243 | 0.852 | 0.933 | - | non coding transcript exon variant | LBX1-AS1 | - |
| rs678741 | 10:101237824 | 18374 | 0.852 | 0.933 | - | intron variant | LBX1-AS1 | - |

**No variants found** Variants linked to rs11190870 in 1000GENOMES:phase_3:CHB

**No variants found** Variants linked to rs11190870 in 1000GENOMES:phase_3:JPT

##### Variants linked to rs11190870 in 1000GENOMES:phase_3:KHV

[back to top]

Show/hide columns Filter

| Variant | Location | Distance (bp) | $r^2$ | D' | Associated phenotype(s) | Consequence Type | Located in gene(s) | Gene phenotype(s) |
|---|---|---|---|---|---|---|---|---|
| rs1407409 | 10:101225650 | 6200 | 0.803 | 1.000 | - | regulatory region variant | - | - |

##### Variants linked to rs11190870 in 1000GENOMES:phase_3:ASW

[back to top]

Show/hide columns Filter

| Variant | Location | Distance (bp) | $r^2$ | D' | Associated phenotype(s) | Consequence Type | Located in gene(s) | Gene phenotype(s) |
|---|---|---|---|---|---|---|---|---|
| rs3950032 | 10:101214352 | 5098 | 0.869 | 1.000 | - | TF binding site | - | - |

| | | | | | | | | |
|---|---|---|---|---|---|---|---|---|
| rs1407409 | 10:101225650 | 6200 | 0.833 | 0.962 | - | regulatory region variant | - | - |
| rs1322330 | 10:101231902 | 12452 | 0.833 | 0.962 | - | intron variant | LBX1-AS1 | - |
| rs625039 | 10:101233892 | 14442 | 0.802 | 0.961 | - | intron variant | LBX1-AS1 | - |

#### Variants linked to rs11190870 in 1000GENOMES:phase_3:LWK

[back to top]

Show/hide columns

Filter

| Variant | Location | Distance (bp) | $r^2$ | D' | Associated phenotype(s) | Consequence Type | Located in gene(s) | Gene phenotype(s) |
|---|---|---|---|---|---|---|---|---|
| rs3950032 | 10:101214352 | 5098 | 0.899 | 0.958 | - | TF binding site | - | - |
| rs1407409 | 10:101225650 | 6200 | 0.861 | 0.957 | - | regulatory region variant | - | - |
| rs1322330 | 10:101231902 | 12452 | 0.800 | 0.895 | - | intron variant | LBX1-AS1 | - |

#### Variants linked to rs11190870 in 1000GENOMES:phase_3:YRI

[back to top]

Show/hide columns

Filter

| Variant | Location | Distance (bp) | $r^2$ | D' | Associated phenotype(s) | Consequence Type | Located in gene(s) | Gene phenotype(s) |
|---|---|---|---|---|---|---|---|---|
| rs3950032 | 10:101214352 | 5098 | 0.958 | 1.000 | - | TF binding site | - | - |
| rs11284675 | 10:101210818 | 8632 | 0.938 | 1.000 | - | intergenic variant | - | - |
| rs10786627 | 10:101212211 | 7239 | 0.919 | 1.000 | - | intergenic variant | - | - |
| rs7913672 | 10:101212938 | 6512 | 0.916 | 0.978 | - | regulatory region variant | - | - |
| rs1407409 | 10:101225650 | 6200 | 0.801 | 0.953 | - | regulatory region variant | - | - |

#### Variants linked to rs11190870 in 1000GENOMES:phase_3:CHS

[back to top]

Show/hide columns

Filter

| Variant | Location | Distance (bp) | $r^2$ | D' | Associated phenotype(s) | Consequence Type | Located in gene(s) | Gene phenotype(s) |
|---|---|---|---|---|---|---|---|---|
| rs1407409 | 10:101225650 | 6200 | 0.857 | 1.000 | - | regulatory region variant | - | - |
| rs1322330 | 10:101231902 | 12452 | 0.857 | 1.000 | - | intron variant | LBX1-AS1 | - |
| rs3950032 | 10:101214352 | 5098 | 0.841 | 1.000 | - | TF binding site | - | - |
| rs143492549 | 10:101222651-101222658 | 3201 | 0.808 | 1.000 | - | intergenic variant | - | - |

#### Variants linked to rs11190870 in 1000GENOMES:phase_3:CDX

[back to top]

Show/hide columns

Filter

| Variant | Location | Distance (bp) | $r^2$ | D' | Associated phenotype(s) | Consequence Type | Located in gene(s) | Gene phenotype(s) |
|---|---|---|---|---|---|---|---|---|
| rs12771674 | 10:101205552 | 13898 | 0.808 | 0.909 | - | intergenic variant | - | - |
| rs11598564 | 10:101204847 | 14603 | 0.808 | 0.909 | - | intergenic variant | - | - |

Ensembl release 106 - Apr 2022 © EMBL-EBI
http://www.ensembl.org
